# Sociodemographic predictors of acute corneal hydrops in patients with unstable keratoconus

**DOI:** 10.1101/2025.01.29.25321351

**Authors:** Levi N Kanu, Nikolay Boychev, Meredith Fry, Leah Margolis, Cherry Le, Rimsha Paneru, Veronica Ng, Jules Hutchison, Joseph B. Ciolino

## Abstract

**Objective:** To determine whether sociodemographic factors affect the outcomes of patients with unstable keratoconus.

**Design:** Retrospective case control study

**Subjects:** All patients diagnosed with unstable keratoconus at Mass Eye and Ear between January 2016 and October 2022. Those who experienced acute corneal hydrops during the study period were considered cases, and those who did not were considered controls.

**Methods:** Potential subjects were identified by billing code and verified with review of clinical documentation. Charts were reviewed for pertinent sociodemographic, medical, and ocular risk factors.

**Main Outcome Measures:** Clinical diagnosis of acute corneal hydrops was the primary outcome. Receipt of corneal crosslinking (CXL) was a secondary outcome.

**Results:** Of 762 patients diagnosed with unstable keratoconus, we identified 128 episodes of hydrops in 113 patients (14.9%). Compared to controls, cases were a significantly higher proportion Black (27.6% vs 14.6%, *P*<0.0001), unemployed (22.4% vs 12.5%, *P*<0.01), and/or covered by public health insurance plans (50.4% vs 27.7%, *P*<0.0001). Body mass index (BMI) was significantly higher in cases (34.1±10.0) than controls (29.2±8.1, mean±SD, *P*<0.0001). Developmental delay (17.7% vs 3.4%, *P*<0.0001), atopy (48.9% vs 30.4%, *P*=0.0001), sleep apnea (9.7% vs 1.2%, *P*<0.0001), and eye rubbing (85.5% vs 67.7%, *P*<0.01) were all more common in the hydrops group. In a stepwise multivariable logistic regression analysis, Black race [2.10 (1.02-4.27), *P*=0.041], public health insurance coverage [2.06 (1.12-3.80), *P*=0.020], BMI [1.05 (1.02-1.08), *P*=0.003], and developmental delay [16.42 (5.62-56.30), *P*<0.001] were each associated with a higher odds of incident hydrops [OR (95%CI)]. Similar trends were found to influence the receipt of CXL, with male sex [0.34 (0.13-0.77), *P*=0.015], Black race [0.39 (0.16-0.97), *P*=0.038], and public health insurance [0.37 (0.16-0.82), *P*=0.014] negatively influencing the odds of receiving CXL.

**Conclusions:** This study establishes several sociodemographic (Black race, unemployment, and publicly insured) and medical (high BMI and developmental delay) factors as independent risk factors influencing the incidence of hydrops and/or the receipt of CXL in a large group of patients with unstable keratoconus. Special attention is warranted when caring for patients within these subpopulations with unstable keratoconus, and further study into the mechanisms of these associations is warranted.

---

Keratoconus is an ectatic corneal disorder with a characteristic pattern of corneal steepening and thinning. Acute corneal hydrops (herein referred to simply as hydrops) is a sudden onset of corneal edema, typically due to a spontaneous break in Descemet membrane that may occur in the setting of ectatic corneal disorders such as keratoconus. Patients with hydrops experience pain, photophobia, and decreased vision at the time of onset, and are often left with chronic, visually significant corneal scarring that frequently necessitates corneal transplantation to improve their vision^1^.

While the break in Descemet membrane precipitating hydrops is typically described as spontaneous, several established risk factors are associated with this condition. Patients with more severe ectasia (i.e., steeper keratometry and worse baseline visual acuity) are more likely to develop hydrops^2–4^. Furthermore, many of the clinical risk factors associated with keratoconus are also associated with the development of hydrops, including male sex^2,3,5^, eye rubbing^5,6^, learning disability^2,4,6^, elevated intraocular pressure^6^, and atopy (e.g. allergic keratoconjunctivitis, asthma, and atopic dermatitis)^2,3^. In addition to clinical and anatomic risk factors, sociodemographic factors like race and ethnicity may influence the risk of developing hydrops in those with keratoconus. For example, in New Zealand, keratoconus patients of Pacific ethnicity have been found to be significantly more likely to develop hydrops than those of European ethnicity^5^. In a study from the United Kingdom, hydrops was significantly more common in patients of South Asian and Black ethnicity than White patients^7^. To our knowledge, a similar evaluation has not yet been performed in the United States. Furthermore, due to the rarity of hydrops—with an estimated overall prevalence in keratoconus patients of less than 3%^3^—studies are often limited by sample size.

Given the well-known associations between sociodemographic factors and keratoconus^8–11^, this study sought to identify potential sociodemographic risk factors associated with the incidence of acute corneal hydrops in a large population of patients diagnosed with unstable keratoconus at an academic center in the United States.

## Methods

This study was approved by the Institutional Review Board at Mass Eye and Ear, with a waiver of the requirement for informed consent. The research was conducted according to the tenets of the Declaration of Helsinki^12^ and in compliance with the Health Insurance Portability and Accountability Act.

This was a retrospective, case-control study of all patients seen at Mass Eye and Ear and diagnosed with unstable keratoconus in one or both eyes between January 1, 2016 and October 31, 2022. Potential study subjects were identified through the Research Patient Data Registry (RPDR), a service provided by Mass General Brigham. All patients receiving a diagnosis of unstable keratoconus based on the International Classification of Diseases 10^th^ revision (ICD) codes *H18*.*62X* were reviewed for inclusion. While unstable keratoconus lacks a consistent definition, subjects were confirmed by clinical documentation (e.g., worsening keratometry or thinning) following identification by ICD code. Eyes with a history of keratoplasty (penetrating keratoplasty or anterior lamellar keratoplasty), keratorefractive surgery (laser-assisted in situ keratomileusis, photorefractive keratectomy, or small incision lenticule extraction), corneal crosslinking (CXL), or intrastromal corneal ring segments prior to the initial study visit were excluded. Charts indicating a clinical diagnosis of stable keratoconus with stable tomographic measurements (i.e., improperly coded as unstable) were excluded. Additionally, those patients with only one single visit during the study period were excluded. The remaining charts were then reviewed for the incidence of hydrops during the study period. Hydrops was diagnosed clinically, defined as the acute onset of corneal edema, and confirmed by chart review. Patient data were collected between October 2022 and November 2023. Data were analyzed between December 2023 and May 2024. Authors had access to the electronic medical records in order to collect data, but only de-identified data were stored using unique codes for analysis.

Demographic information available through the RPDR included age, sex, reported race and ethnicity, address of residence, health insurance provider, employment status, and preferred language. Address of residence was used to determine national area deprivation index (ADI) using a publicly available dataset through Columbia University^13^. Additionally, all charts were reviewed for medical history. Body mass index was recorded, along with the presence of atopic conditions, developmental delay (including Trisomy 21 and cerebral palsy), sleep apnea, connective tissue disorders (including Ehlers Danlos Syndrome and Osteogenesis imperfecta), reported history of eye rubbing, contact lens wear, and comorbid ocular disease. Visual acuity and biometric data from the latest visit during the study period prior to a hydrops event were recorded, including the mean and maximum keratometry readings, anterior astigmatism, and thinnest cornea reading. Surgical outcomes (i.e., CXL or keratoplasty), and latest follow-up were recorded for all patients.

Comparisons between the sociodemographic, medical, and ocular characteristics of case and control populations were made using the Student’s *t*-test, χ^2^ test, and Fisher’s exact test, as appropriate. Associations of these factors and the development of hydrops were assessed using a multivariate logistic regression model. Considering the number of predictors analyzed, a stepwise regression analysis was also performed. Patients with missing information for one or more of the tested variables were excluded from the regression analyses. Separately, subjects who did not experience hydrops were included in a similar logistic regression analysis of factors influencing receipt of CXL.

## Results

Of 1076 patients screened for inclusion, 314 were excluded based on the outlined inclusion and exclusion criteria. Of the remaining 762 subjects, we identified 128 episodes of hydrops in 113 patients (14.9%) during the study period, while 1118 eyes with unstable keratoconus of the remaining 649 patients (88.3%) did not experience hydrops (controls). Each group comprised a wide age range (10-69 hydrops; 11-74 controls). On average, hydrops subjects were slightly older at the time of incident hydrops than controls at their latest follow-up. Eight patients experienced hydrops in both eyes, and two patients experienced consecutive episodes of hydrops in the same eye. One patient presented with hydrops in both eyes simultaneously, while the average interval between episodes of consecutive hydrops was 2.5±2.5 years.

Seven (5.5%) of the eyes experiencing hydrops patients had undergone CXL prior to their hydrops event. Of these, the majority (71.4%) were transepithelial CXL. In those post-CXL patients who experienced hydrops, the interval between CXL and hydrops ranged from 6 days to 4 years, with a median interval of 1.25 years (IQR 1.8 months-2.5 years).

In addition to medical management, 39 surgical procedures were performed on 32 eyes for the management of hydrops or its sequelae, including intracameral gas injection (n=14) and keratoplasty (either endothelial keratoplasty or penetrating keratoplasty, n=20). Seven eyes underwent CXL following resolution of hydrops.

Forty-two (37.2%) of the patients with hydrops had established care within one of the Mass Eye and Ear clinics prior to the hydrops event. The remaining 71 patients with hydrops saw a Mass Eye and Ear provider for the first time after their hydrops event. Nine patients (8.0%) were lost to follow-up after their initial evaluation for hydrops. Of those hydrops patients not lost to follow-up, median follow-up was 3.3 years (IQR 1.2-5.2). In comparison, controls had a median follow-up of 11 months (IQR 0.3-2.0 years). A majority (92.1%) of patients who did not experience hydrops during the study period underwent CXL, after which they were excluded from further analysis in this study.

Hydrops disproportionately affected those of Black race, those who were not employed, and those with public healthcare plans (Table 1). Developmental delay, atopy, sleep apnea, and a self-reported history of eye rubbing were all disproportionately represented in the hydrops group (Table 1). Additionally, those with hydrops tended to have a higher body mass index (BMI). Contact lens use was more common among those with hydrops, but this difference was not statistically significant. Baseline tomography was worse in those who experienced hydrops (Figure 1). Specifically, mean keratometry (64.5±11.2 vs 50.0±8.3, p<0.0001) and maximum keratometry values (79.4±13.1 vs 59.0±12.7, p<0.0001) were higher in those with hydrops compared to controls, while thinnest corneal thickness values (328.3±96.1 vs 456.2±59.1, p<0.0001) were lower. Subjects with Black race, non-employed status, public health insurance coverage were disproportionately affected by hydrops. While cases tended to live in areas of higher disadvantage (i.e., higher ADI), this difference was not significant. To determine the independent effect of sociodemographic factors on risk of hydrops, multivariate logistic regression modeling was performed using predictors for the 337 subjects (80 cases and 257 controls) who had all predictor variables known (Table 2).

**Figure 1.**
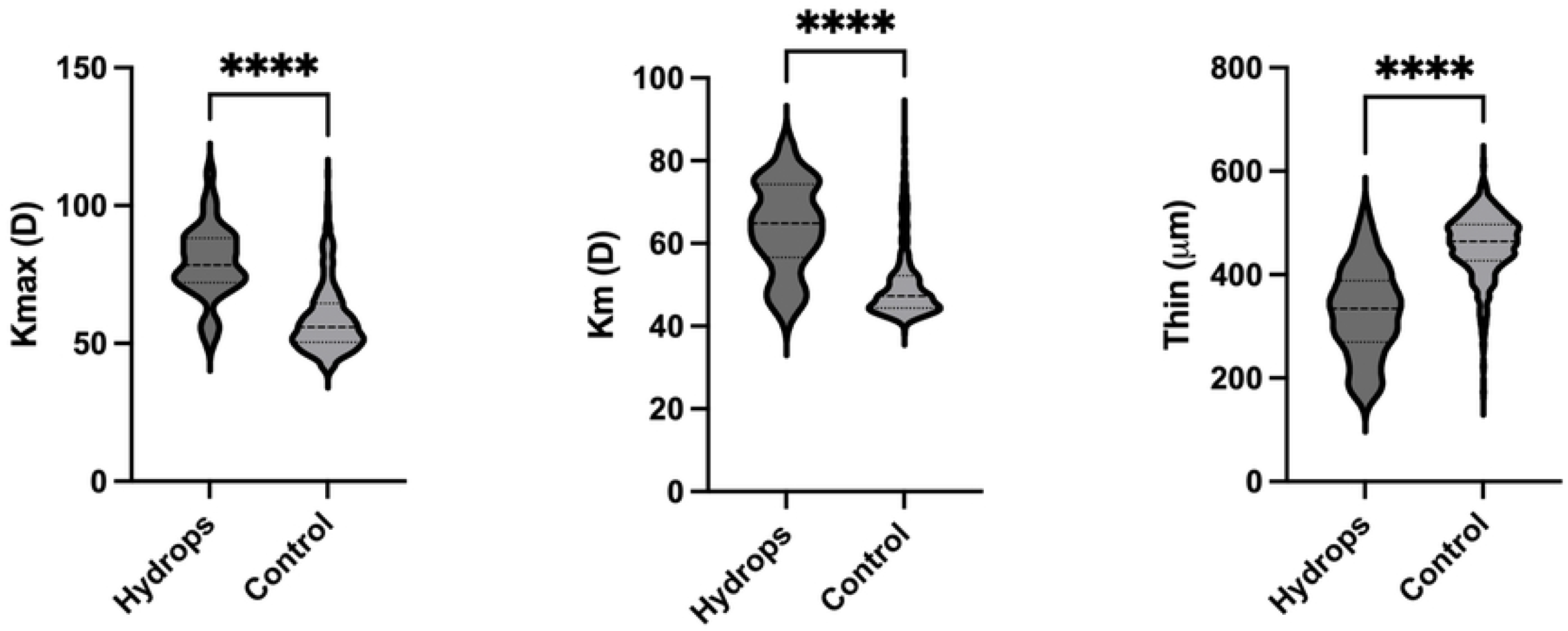
Baseline tomographic values in subjects with unstable keratoconus who did (*Hydrops*) or did not (*Control*) experience hydrops during the study period. Maximum keratometry (*Kmax*), mean keratometry (*Km*), and thinnest pachymetry (*Thin*) values from the latest tomographic study within the study period are shown (prior to a hydrops event, if applicable). \*\*\*\**P*<0.0001.

A large proportion (92.1%) of those seen with a diagnosis of unstable keratoconus underwent CXL during the study period. Those who did not receive CXL were more likely to be of Black race, have higher no-show rate, have public insurance, and live in an area of higher (worse) area deprivation index (Table 3). Multivariable logistic regression analysis was performed on the subjects who had all predictor variables known (39 who did not receive CXL and 444 who did, Table 4). This analysis revealed race, sex, and insurance as significant independent predictors of CXL.

## Discussion

Our study demonstrates that significant sociodemographic disparities may exist in the outcomes of patients with unstable keratoconus. Specifically, in a stepwise logistic regression model, Black race and public insurance plans were associated with higher odds of experiencing acute corneal hydrops. Similar trends were found in the factors influencing receipt of CXL, with Black race, public insurance plans, and male sex each independently reducing the odds of receiving CXL. Overall, our findings align with a recent study which found sex, race, language, insurance, and employment to be significantly associated with either KCN severity, progression, or a combination thereof^14^.

Our study uncovered a series of eight patients who experienced corneal hydrops in both eyes, including two patients with multiple episodes of hydrops in the same eye. Both patients who had multiple episodes in the same eye also experienced hydrops in the fellow eye. There was no readily identifiable trend associated with this subset of patients experiencing bilateral or sequential hydrops.

In accordance with most published evidence^2,7^, the majority of patients with unstable keratoconus and hydrops were male. However, our analysis suggests that male sex is not an independent factor predisposing to hydrops in those with unstable keratoconus. Male sex did appear to independently influence the receipt of CXL, suggesting that the effects of sex were sociodemographic rather than biological in nature. Indeed, while mechanisms are still being elucidated, male patients exhibit lower overall healthcare and preventive care utilization on a population level^15,16^. Similarly, Black race in particular has long been associated with reduced access to and underutilization of healthcare systems^17,18^, which may explain the associations seen in our study. Interestingly, while employment characteristics were significantly different between the case and control groups, these factors were not significant in our multivariate logistic regression models of hydrops and CXL. This suggests a correlation between employment status and other measured variables, including race and health care insurance. Additionally, while ADI was higher among those with hydrops, this difference was not statistically significant, and multivariate analysis did not reveal ADI as an independent risk factor. Given that our patient population included patients from out-of-state, a national ranking rather than state ranking was used in ADI. However, since the vast majority of residential areas within the state of Massachusetts are within the top three deciles nationally^19,20^, we suspect that ADI differences were minimized with our methodology.

While the U.S. Food and Drug Administration approved corneal crosslinking for keratoconus in 2016^21^, insurance coverage of the procedure has been variable—albeit increasing—since then. Even with insurance coverage, out-of-pocket costs and indirect costs (i.e., lost wages for those who may be self-employed or not have paid sick time) may still be substantial and affect utilization^22^. Our analysis demonstrated that patients covered by public healthcare plans (i.e., Medicare and Medicaid) were more likely to experience hydrops and less likely to undergo CXL during the study period. Whether this association is due to differences in coverage for the procedure, out-of-pocket expenses, or confounding with other variables remains to be elucidated. The uninsured group was highly represented in the hydrops group, but the low number of uninsured patients meant our study was not powered to detect a significant difference.

Our findings also raise several other important considerations regarding CXL. Despite being highly effective in slowing the progression of KCN^23^, tomographic progression may still occur following CXL in a subset of patients^24,25^. It follows that the risk of hydrops may not be entirely mitigated by CXL, and—while rare—cases of hydrops following CXL have been reported^26,27^. In our study, while the overall incidence of hydrops in post-CXL patients was low (1.2%), a relatively high percentage (6.2%) of hydrops patients had a history of CXL, and a relatively high proportion (71.4%) of these were trans-epithelial CXL. We suspect that those receiving trans-epithelial CXL may be more likely to have severe disease (i.e., pachymetry too thin for epi-off CXL^28^) and thus more likely to experience hydrops, but this association warrants further investigation. These findings also reinforce the need to continue to follow patients for progression following CXL.

Of the systemic medical conditions evaluated, developmental delay (including trisomy 21 and cerebral palsy) and elevated BMI were found to significantly increase the odds of experiencing hydrops. While the average BMI of patients with unstable keratoconus was already in the obese range, our results indicate a strongly positive correlation between BMI and hydrops incidence within this population. Multiple recent cross-sectional or case control studies have demonstrated a significant association between BMI and keratoconus^29,30^, and recent evidence of a causal relationship between obesity and keratoconus has been established^31^. Our study provides further support to this connection between obesity and keratoconus.

### Limitations

The study has several limitations. As a study confined to a single institution, our findings may not be generalizable to other populations. The study was limited by its retrospective design, including the limitations of quality and completeness of data. Several factors were self-reported (e.g., race and ethnicity), and these factors were omitted in a substantial proportion of patients. Many patients were self-reported as “other” race which, given the diversity of our patient population, likely indicates that this group is quite diverse racially. Eye rubbing and contact lens wear were also patient-provided data and may have been omitted from some charts. Additionally, as routine height and weight measurements are not often performed in the eye clinic, factors such as BMI were unavailable for a significant portion of patients. Data incompleteness appeared to affect the non-hydrops group more than the hydrops group, which may have introduced bias to the results. Subjects in this study were identified by diagnosis billing codes and, while efforts were made to verify these with medical record analysis, subjects improperly coded may have been omitted from the initial study set. We could not account for patients who were seen for only one single visit, and this may have introduced bias in our findings. Additionally, differential loss to follow-up may have influenced our findings.

## Conclusions

In conclusion, our study has identified several sociodemographic and systemic medical conditions that affect outcomes in patients with unstable keratoconus. Black race, public health insurance coverage, obesity, and developmental delay each independently increased the odds of hydrops in our population. Moreover, male sex, Black race, and public health insurance coverage each independently reduced the odds of receiving CXL in those with unstable keratoconus at our institution. Further study into the mechanisms of each of these associations is warranted. These findings may guide the follow-up and treatment recommendations of clinicians caring for patients with unstable keratoconus.

## Data Availability

All relevant data are within the manuscript and its Supporting Information files.

